# Daily heart rate variability biofeedback training decreases locus coeruleus MRI contrast in younger adults

**DOI:** 10.1101/2022.02.04.22270468

**Authors:** Shelby L. Bachman, Steve Cole, Hyun Joo Yoo, Kaoru Nashiro, Jungwon Min, Noah Mercer, Padideh Nasseri, Julian F. Thayer, Paul Lehrer, Mara Mather

## Abstract

As an arousal hub region in the brain, the locus coeruleus (LC) has bidirectional connections with the autonomic nervous system. Magnetic resonance imaging (MRI)-based measures of LC structural integrity have been linked to cognition and arousal, but less is known about factors that influence LC structure and function across time. Here, we tested the effects of heart rate variability (HRV) biofeedback, an intervention targeting the autonomic nervous system, on LC MRI contrast and sympathetic activity. Younger and older participants completed daily HRV biofeedback training for five weeks. Those assigned to an experimental condition performed biofeedback involving slow, paced breathing designed to increase heart rate oscillations, whereas those assigned to a control condition performed biofeedback to decrease heart rate oscillations. At the pre- and post-training timepoints, LC contrast was assessed using turbo spin echo MRI scans, and RNA sequencing was used to assess cAMP-responsive element binding protein (CREB)-regulated gene expression in circulating blood cells, an index of sympathetic nervous system signaling. We found that left LC contrast decreased in younger participants in the experimental group, and across younger participants, decreases in left LC contrast were related to the extent to which participants increased their heart rate oscillations during training. Furthermore, decreases in left LC contrast were associated with decreased expression of CREB-associated gene transcripts. On the contrary, there were no effects of biofeedback on LC contrast among older participants in the experimental group. These findings provide novel evidence that in younger adults, HRV biofeedback involving slow, paced breathing can decrease both LC contrast and sympathetic nervous system signaling.

## 1 Introduction

The locus coeruleus (LC), a small nucleus in the brainstem, helps coordinate the brain’s arousal system. Situated at the lateral floor of the fourth ventricle, the LC serves as the brain’s primary source of the neurotransmitter norepinephrine (NE; Schwarz & Luo, 2015). NE released from the LC to the brain and spinal cord regulates wakefulness, coordinates adaptive behavior, and modulates processes of learning and memory (Berridge & Waterhouse, 2003; Sara, 2009). As an arousal center, the LC is involved in the central stress response (Koob, 1999): During acute stress, corticotropin releasing factor released onto the LC promotes elevated levels of tonic LC neuronal activity, thereby facilitating cortical NE release, the peripheral sympathetic fight-or-flight response and anxiety-like behaviors (Curtis et al., 1997; McCall et al., 2015; Valentino & Van Bockstaele, 2008). Experienced over the longer term, stress can have maladaptive effects on LC function and structure (Morris et al., 2020), with chronic corticotropin releasing factor exposure being linked to increased LC neuronal sensitivity, firing rates, and dendritic arborization in rodents (Borodovitsyna et al., 2018). Yet the LC has not only been implicated in arousal and stress: It is also the first brain location where tau pathology accumulates in the progression of Alzheimer’s disease (Braak et al., 2011). LC neurodegeneration is characteristic of Alzheimer’s disease (Chalermpalanupap et al., 2017), and older adults with relatively lower cell density within the LC exhibit faster rates of cognitive decline prior to death (Wilson et al., 2013).

In recent years, specialized magnetic resonance imaging (MRI) sequences have accelerated study of the human LC via their ability to quantify LC structure in vivo (Sasaki et al., 2006). In these sequences, the LC exhibits elevated MRI signal contrast relative to surrounding tissue, and LC MRI contrast is thought to reflect LC structural integrity (Betts et al., 2019). Across studies, having higher LC contrast has been linked to better cognitive outcomes in older adults (Dahl et al., 2019; Hämmerer et al., 2018; Liu et al., 2020), as well as reduced risk of developing mild cognitive impairment (Elman et al., 2021) and fewer preclinical Alzheimer’s disease processes (Jacobs et al., 2021). We recently reported that contrast of the rostral LC was positively associated with cortical thickness in various brain regions in older adults, whereas caudal LC contrast showed some negative associations with cortical thickness in younger adults (Bachman et al., 2021). Thus, higher LC contrast may not always be a positive indicator. For instance, one study found that LC volume - also quantified using an MRI sequence that yields elevated signal intensity in the LC - was positively correlated with anxious arousal and self-reported general distress in younger adults (Morris et al., 2020). Likewise, another study found that participants with relatively higher LC contrast had lower heart rate variability (Mather et al., 2017). These findings suggest that tonic high noradrenergic activity associated with stress, or a relative dominance of sympathetic over parasympathetic activity, are also associated with high LC contrast in younger adults. Our hypothesis for these seemingly discrepant findings in younger and older adults is that different factors contribute to LC contrast across the lifespan: Earlier in life, stress is the primary factor influencing individual differences in LC contrast, whereas later in life, neurodegeneration is the dominant factor influencing individual differences in LC contrast.

Yet despite what has been learned about the LC from recent MRI studies, no published studies have assessed whether LC MRI contrast can be changed. What might give rise to changes in LC contrast over time? Because the LC is a key player in the stress response and has bidirectional connections with the parasympathetic and sympathetic branches of the autonomic nervous system (Wood et al., 2017), we reasoned that an intervention targeting the autonomic nervous system could influence both LC structure and function. One such intervention is heart rate variability (HRV) biofeedback (Lehrer & Gevirtz, 2014). Individuals with higher HRV, an index of parasympathetic control over heart rate and autonomic regulation (Mulcahy et al., 2019; Thayer & Lane, 2000), are better able to regulate their emotions and exhibit reduced physiological responses to stressors (Thayer & Lane, 2009; Weber et al., 2010), relative to those with lower HRV. HRV can be systematically manipulated through biofeedback that involves slow, paced breathing and simultaneous feedback on the coupling between heart rate oscillations and breathing (Lehrer & Gevirtz, 2014). Slow breathing, particularly at a pace around 10 seconds per breath, elicits maximally high-amplitude oscillations in heart rate (Lehrer et al., 2003). Slow breathing also stimulates the vagus nerve (Brown & Gerbarg, 2005), which sends projections to the LC by way of the nucleus tractus solitarii (NTS; Badran et al., 2018; Fornai et al., 2011). Performing HRV biofeedback over a period of weeks has been shown to reduce levels of stress and anxiety (Goessl et al., 2017) in younger as well as older adults (Jester et al., 2019), but it is unknown whether HRV-biofeedback affects the LC’s structure and function.

Here, we examined whether HRV biofeedback affected LC MRI contrast and sympathetic activity. Younger and older participants completed 5 weeks of HRV biofeedback training as part of a clinical trial testing the effects of HRV biofeedback training on brain regions involved in emotion regulation (Clinicaltrials.gov NCT03458910 “Heart Rate Variability and Emotion Regulation,” Nashiro et al., 2021). Participants in an experimental condition completed daily biofeedback involving slow, paced breathing to increase heart rate oscillations and HRV, whereas participants in an active control condition completed daily biofeedback training designed to decrease heart rate oscillations and HRV. Both before and after the 5-week training period, we assessed LC contrast in all participants using turbo spin echo (TSE) MRI scans that exhibit elevated signal intensity in the LC. Based on prior work demonstrating beneficial effects of HRV on emotional well-being, and in line with our hypotheses regarding LC contrast reflecting stress in younger adults, we expected that performing 5 weeks of HRV biofeedback training would decrease LC contrast in younger participants. Conversely, as we hypothesize that neurodegeneration, more so than stress, shapes LC contrast in older adulthood, we predicted that HRV biofeedback would either not change or would increase LC contrast in older participants. Furthermore, in a subset of younger participants, we collected blood samples before and after the training period to assess changes in a health-relevant index of sympathetic nervous system (SNS) activity – blood cell expression of genes regulated by the cAMP-responsive element binding protein (CREB) family of transcription factors, which mediates beta-adrenergic signaling from the SNS (Cole et al., 2010; Mayr & Montminy, 2001). In this subset, we predicted that training-related decreases in LC contrast would be coupled with decreases in SNS signaling and thereby reduce expression of CREB-regulated gene transcripts. Previous research has validated blood cell CREB-associated RNA expression levels as a measure of beta-adrenergic signaling (Brown et al., 2010; Cole et al., 2005; Powell et al., 2013).

## 2 Methods

### 2.1 Participants

Data were collected as part of an intervention study testing the effects of 5 weeks of HRV biofeedback training in younger and older adults (for a full description of the study, see Nashiro et al., 2021). Participants in the study were assigned to one of two conditions. Those in an increase-oscillations (Osc+) condition completed 20-40 minutes of daily biofeedback training involving slow, paced breathing which was designed to increase heart rate oscillations and HRV. Participants in a decrease-oscillations (Osc-) condition completed 20-40 minutes of biofeedback training per day designed to decrease heart rate oscillations and HRV. Eligible participants were healthy, MRI-eligible younger and older adults recruited from the University of Southern California and Los Angeles communities. Individuals who regularly practiced biofeedback training or breathing techniques were excluded from participation. Older adults were screened for cognitive dysfunction by telephone using the TELE interview (Gatz et al., 1995); individuals scoring below 16 were excluded from participation.

As part of the intervention study which lasted 7 weeks, MRI assessments were conducted at a pre-training timepoint (second study week), before participants learned about or practiced the intervention, and following 5 weeks of biofeedback training (seventh study week). A total of 175 participants (115 younger, 60 older) completed pre- and/or post-training MRI assessments, yielding a total of 325 TSE scans (detailed breakdown in the Supplementary Methods, Section 1). Following exclusions for artifact or motion on native TSE scans (Section 2.3.1), 287 scans were used for LC delineation. Additional exclusions were applied due to artifact after warping TSE scans to MNI152 space (Section 2.3.1). In addition, blood samples were collected from a subset of 54 younger participants at pre- and post-training timepoints (first and sixth study weeks, respectively) to assess change in expression of genes regulated by CREB. A total of 129 participants (93 younger, 36 older) with LC contrast values and/or blood-based measures available at both timepoints were included for analysis. Characteristics of this sample are presented in Table 1. The University of Southern California Institutional Review Board approved the study. All participants provided written, informed consent prior to participation and received monetary compensation for their participation.

**Table 1:** Sample characteristics.

| Age group | Condition | N | N (%)<br>Female | Age, mean<br>(SD) | Age,<br>range | Education,<br>mean (SD) | Education,<br>range |
| --- | --- | --- | --- | --- | --- | --- | --- |
| Younger | Osc+ | 47 | 26 (55.3%) | 22.64 (2.57) | 18-28 | 15.99 (1.87) | 12-20 |
| Younger | Osc- | 46 | 24 (52.2%) | 22.57 (3.24) | 18-31 | 15.74 (2.64) | 12-24 |
| Older | Osc+ | 17 | 12 (70.6%) | 65 (6.86) | 55-80 | 17.12 (2.71) | 14-25 |
| Older | Osc- | 19 | 15 (78.9%) | 65.21 (5.61) | 57-77 | 16.53 (2.39) | 14-22 |

*Note.* Age and education are expressed in years. Osc+ = increase-oscillations condition; Osc- = decrease-oscillations condition; SD = standard deviation.

### 2.2 MRI data collection

MRI data were collected at the University of Southern California David and Dana Dornsife Neuroimaging Center, on a Siemens Magnetom Trio 3T MRI scanner with a 32-channel head coil. Sequences relevant to the present analyses are described below.

A high-resolution, T_1_-weighted magnetization prepared rapid acquisition gradient echo (MPRAGE) scan was acquired (TR = 2300ms, TE = 2.26 ms, flip angle = 9°, bandwidth = 200 Hz/Px, isometric voxel size = 1.0mm^3^, no gap between slices, 175 volumes). Based on the MPRAGE scan, a two-dimensional, multi-slice TSE scan was collected by aligning the field of view perpendicular to the respective participant’s brainstem. Parameters of this TSE sequence were as follows: TR = 750ms, TE = 12ms, flip angle = 120°, bandwidth = 287 Hz/Px, voxel size = 0.43 x 0.43 x 2.5mm, gap between slices = 1mm. The TSE sequence included 11 axial slices and covered the entire pons. TSE scans from randomly selected participants are shown in Figure 1A.

**Figure 1.**
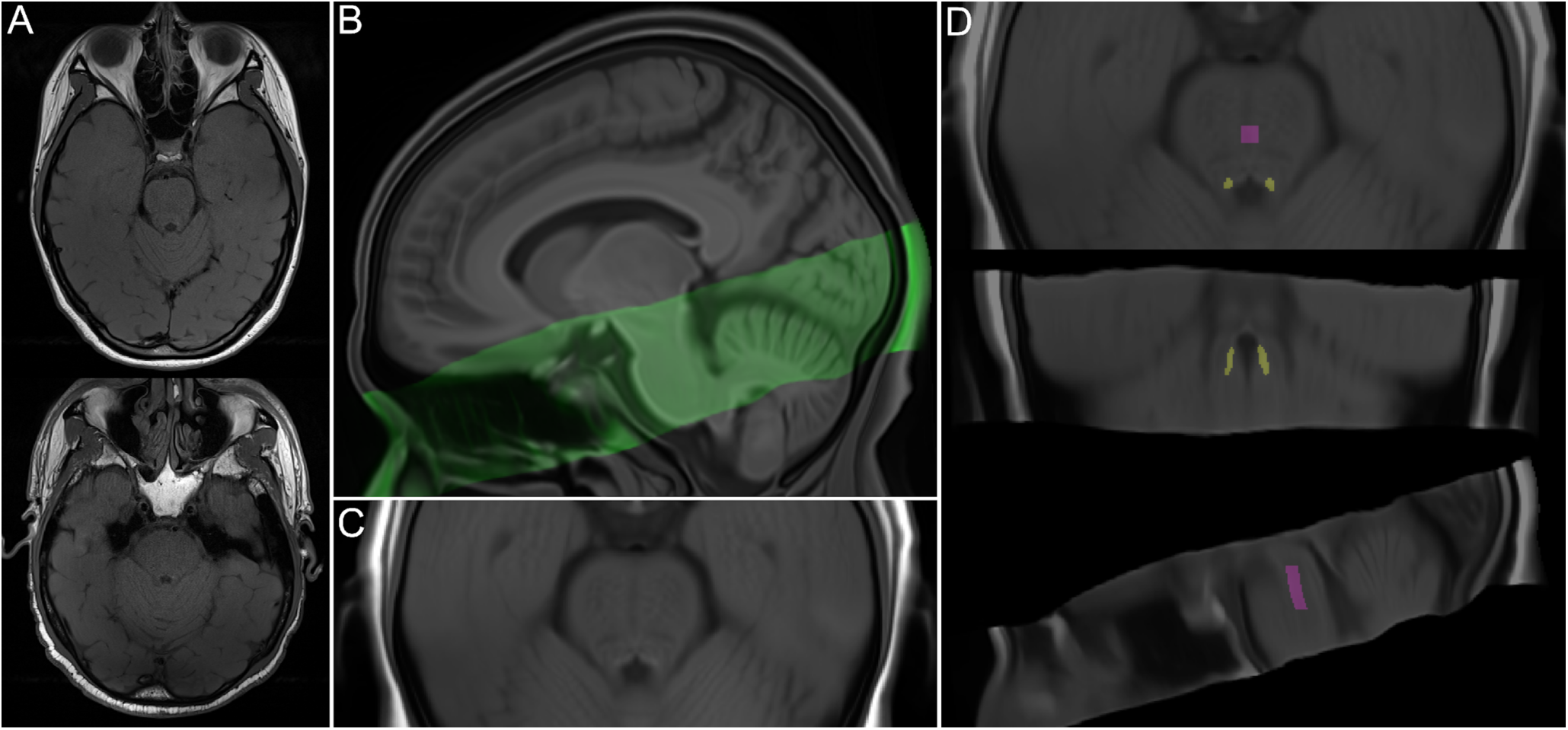
(A) Turbo spin echo (TSE) scans from randomly selected younger (top) and older (bottom) participants. (B) Sagittal view of TSE template (green) overlaid onto whole-brain template, both warped to MNI152 0.5mm (linear) space. (C) Detailed axial view of TSE template, warped to MNI152 space. (D) TSE template, warped to MNI152 space, overlaid with locus coeruleus meta-mask and pontine reference region from Dahl et al. (2021), which were used for calculation of LC contrast ratios.

### 2.3 MRI data analysis

#### 2.3.1 LC delineation

We used a semi-automated method to delineate the LC on all available pre- and post-training TSE scans based on approaches described by Dahl et al. (2019) and Ye et al. (2021). LC delineation steps were performed using Advanced Normalization Tools (ANTs; Version 2.3.4; Avants et al., 2011; http://stnava.github.io/ANTs/). Visualization steps were performed using ITK-SNAP (Version 3.8.0; Yushkevich et al., 2006; http://www.itksnap.org). Parameters for each step are described in the Supplementary Methods (Section 2).

All TSE scans were first visually inspected; scans with excessive motion or susceptibility artifact overlapping the LC or pons (n = 34), incorrect positioning (n = 3), or different resolution (n = 1) were excluded from LC delineation (Supplementary Methods, Section 1). The remaining TSE and corresponding MPRAGE scans were upsampled to twice their native resolution using the ResampleImage ANTs routine. Upsampled MPRAGE scans were used to generate a whole-brain template with the antsMultivariateTemplateConstruction.sh routine (Figure 1B; see Supplementary Methods Section 2 for a description of template-building procedures). Each TSE scan was then coregistered to its corresponding whole-brain template-coregistered MPRAGE scan, using the antsRegistrationSyNQuick.sh routine. All coregistered TSE scans were used to build a TSE template (Figures 1B and 1C). Using the antsRegistrationSyN.sh routine, the resulting TSE template was coregistered to the whole-brain template to ensure spatial alignment. The whole-brain template was then coregistered to MNI152 0.5mm (linear) standard space, in order to facilitate comparison with previously-published LC maps. Transforms from all template-building and coregistration steps described above were applied in a single step to warp upsampled TSE scans to MNI152 space, using the antsApplyTransforms.sh routine. In addition, transforms from the final coregistration steps were applied to warp the TSE template to MNI152 space (Figure 1C). As a validation step, we examined whether locations of hyperintensity on the TSE template in MNI152 space aligned with the location of the Dahl et al. (2021) meta-map, which was generated by aggregating across published LC maps and thus reflected a plausible LC volume of interest with high agreement across studies. We found high correspondence between hyperintensities on the TSE template and the LC meta-map (Supplementary Figure S1).

At this stage, a total of 6 warped TSE scans were excluded from LC delineation after visually confirming that, once warped to MNI152 space, they contained artifacts overlapping the LC or central pons. This left data from 78 younger (39 Osc+, 39 Osc-) and 36 older (17 Osc+, 19 Osc-) participants included for LC delineation and analyses of change in LC contrast. We proceeded to delineate the LC for individual participants and timepoints by applying the Dahl et al. (2021) LC meta-map as a mask on all warped TSE scans (Figure 1D). Within the masked region of each scan, we extracted the intensity and location of the peak-intensity LC voxel in each *z*-slice and hemisphere. As another validation step, we compared the resulting intensity values to intensity values determined through manual delineation of the LC on native-resolution TSE scans (Supplementary Methods, Section 3). Two-way mixed-effects intraclass correlation analyses indicated high correspondence between peak LC intensity values from the semi-automated and manual methods for the left LC (*ICC*(C, 1) = 0.939, *95% CI* = 0.921 - 0.953, *p* < .001) and right LC (*ICC*(C, 1) = 0.924, *95% CI* = 0.902 - 0.941, *p* < .001). To compute LC contrast ratios reflecting peak LC intensity relative to that of surrounding tissue, we also extracted intensity values from a central pontine region (Figure 1D). Specifically, we applied the central pontine reference map from Dahl et al. (2021) as a mask on individual TSE scans that had been warped to MNI space and extracted the peak intensity value within the masked region.

#### 2.3.2 Calculation of LC MRI contrast

LC MRI contrast is typically calculated as a ratio reflecting peak signal intensity in the LC relative to peak intensity within a pontine reference region (Liu et al., 2017):

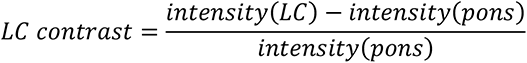

No published studies have examined the stability of peak LC signal intensity locations across time, or factors which influence locations of peak LC intensity, thus we performed an exploratory step to guide our calculation of LC contrast. Specifically, we assessed whether, for each participant, locations of peak intensity in each left and right LC shifted from pre- to post-training (peak LC intensity locations are depicted in Supplementary Figure S2A). To do so, we calculated for each participant the 3-dimensional distance between peak LC intensity locations at the pre- and post-training timepoints, for left and right LC separately (Supplementary Figure S2B). A linear mixed effects analysis indicated that these distances differed from 0 across training conditions, age groups and hemispheres (*p* <. 001; Supplementary Results, Section 1), suggesting that locations of peak LC intensity were not consistent within individuals across time. We therefore aimed to calculate LC contrast in a way that was not biased by peak LC signal intensity location at either the pre- or post-training timepoint. Specifically, for each participant, we calculated LC contrast at each timepoint as an average of LC contrast at the locations of pre- and post-training peak LC signal intensity.

### 2.4 Blood sampling and RNA sequencing analysis

For a subset of participants (N = 54 younger adults), peripheral blood samples were collected under resting conditions at the pre- and post-training timepoints by antecubital venipuncture into PAXgene RNA tubes. Following collection, samples were gently inverted ten times and kept at room temperature for between 1.48 and 22.20 hours (mean = 3.39 hours). Samples were then stored frozen at -80°C at the USC School of Gerontology before they were transferred and assayed in a single batch at the UCLA Social Genomics Core Laboratory, as previously described (Cole et al., 2020). Briefly, total RNA was extracted from 2.5 ml blood samples using an automated nucleic acid processing system (QIAcube; Qiagen), checked for suitable RNA integrity and mass (>50 ng by NanoDrop One spectrophotometry; achieved mean = 4497 ng) and assayed by RNA sequencing in the UCLA Neuroscience Genomics Core Laboratory using Lexogen QuantSeq 3’ FWD cDNA library synthesis and multiplex DNA sequencing on an Illumina HiSeq 4000 instrument with single-strand 65-nt sequence reads (all following the manufacturer’s standard protocol). Analyses targeted >10 million sequence reads per sample (achieved mean 15.1 million), each of which was mapped to the RefSeq human transcriptome sequence using the STAR aligner (achieved average 94% mapping rate) to generate transcript counts per million total transcripts (TPM). TPM values were floored at 1 TPM to reduce spurious variability, log2-transformed to reduce heteroscedasticity, and analyzed by linear statistical models with promoter sequence-based bioinformatics analyses of CREB activity as described below.

### 2.5 Statistical analysis

We fit a linear mixed effects model to assess the fixed effects of timepoint, training condition, age group and hemisphere on LC contrast. Mixed models were also fit for each age group separately to examine the fixed effects of timepoint, training condition, hemisphere and their interactions on LC contrast. Significant timepoint x condition interactions were supplemented with post hoc comparisons of pre- versus post-training LC contrast for each training condition and hemisphere.

Next, we tested whether changes in LC contrast were related to the extent to which participants increased their heart rate oscillations during biofeedback training. For each participant, values of change in left and right LC contrast were calculated as the difference between pre- and post-training LC contrast values. As a measure of how much participants increased heart rate oscillations during practice, we calculated a value of training oscillatory power using each participant’s pulse data collected during training sessions (Supplementary Methods, Section 4). We then fit another mixed model testing the fixed effects of training oscillatory power, hemisphere, age group, and their interactions on LC contrast. For each age group and hemisphere separately, we also performed planned Pearson correlation analyses to test associations between change in LC contrast and training oscillatory power.

Based on previous findings of sex differences in LC contrast (Bachman et al., 2021) as well as sex differences in the responsiveness of the LC to stress (Bangasser et al., 2016), we tested for sex differences in LC contrast change and its relationship with training oscillatory power by fitting the previously described mixed models including sex and its interactions as fixed effects (Supplementary Methods, Section 5). These analyses were performed only for younger participants because we were underpowered to detect sex differences among older participants.

These analyses were performed in R (Version 4.1.0; R Core Team, 2021). Linear mixed effects models were fit using the R package ‘lme4’ (Version 1.1-27.1; Bates et al., 2015), and significance of fixed effects was assessed with Satterthwaite’s method as implemented in the R package ‘lmerTest’ (Version 3.1-3; Kuznetsova et al., 2017). All models included random intercepts for participants. A sum coding contrast scheme was applied to factor variables (condition: Osc+ = 0.5, Osc- = 0.5; timepoint: post-training = 0.5, pre-training = 0.5; age group: older = 0.5, younger = -0.5; hemisphere: left = 0.5, right = -0.5; sex: female = 0.5; younger = 0.5). Post hoc comparisons of model-estimated marginal means were performed with the ‘emmeans’ R package (Version 1.6.2-1; Lenth, 2021). Effect sizes were calculated using the R package ‘effectsiz’ (Version 0.5; Ben-Shachar et al., 2020) and reported as *d*.

We also tested whether training condition (Osc+ or Osc-) or change in LC contrast was associated with change in CREB activity from pre- to post-training using an established bioinformatic measure of CREB gene regulation employed in previous research (Cole et al., 2020). Data from 54 younger participants (30 Osc+, 24 Osc-) with available blood-based measures at both timepoints were included for analysis of CREB activity change by training condition, and data from 39 younger participants (22 Osc+, 17 Osc-) with available blood-based measures and LC contrast values at both timepoints were included for analysis of associations between CREB activity change and LC contrast change. In these analyses, whole transcriptome profiling data were screened to identify genes that showed > 1.5-fold differential change over time between conditions or > 1.5-fold differential change in expression per standard deviation (SD) of pre- to post-training LC contrast change, and the core promoter DNA sequences of those genes were scanned for the prevalence of CREB-binding motifs using the TELiS database (Cole et al., 2020; Cole et al., 2005). Analyses were conducted as previously described (Cole et al., 2020), with CREB activity quantified by the ratio of CREB-binding site prevalence (defined by TRANSFAC position-specific weight matrix V$CREB_Q4) in genes up-regulated in association with condition differences in change or LC contrast change (i.e., >1.5-fold upregulation from pre- to post-training timepoint per SD of LC contrast change) vs. down-regulated (>1.5-fold down-regulated), and log2-ratios averaged over 9 parametric combinations of promoter sequence length (−300, −600, and −1000 to +200 bp relative to the RefSeq transcription start site) and detection stringency (TRANSFAC mat_sim = .80, .90, and .95). Statistical significance was assessed using standard errors derived from bootstrap resampling of linear model residual vectors in underlying gene expression data, which controls for correlation across genes. For additional details on analytic methods, see Cole et al. (2005) and Cole et al. (2020).

## 3 Results

### 3.1 LC contrast decreased in younger participants in the Osc+ condition

LC contrast at the pre- and post-training timepoints is shown in Figure 2A. Using a linear mixed effects analysis testing the fixed effects of condition, timepoint and hemisphere on LC contrast in younger participants (Table 2A), we found a significant training condition x timepoint interaction on LC contrast (*p* = .034, *d* = -0.283). Post hoc comparisons of estimated marginal means indicated that at the post-relative to the pre-training timepoint, LC contrast was numerically lower among younger participants in the Osc+ condition (left: *t*(228) = -2.193, *p* = .029, *d* = -0.497; right: *t*(228) = -0.059, *p* = .953, *d* = -0.013) and numerically higher among younger participants in the Osc-condition (left: *t*(228) = 0.599, *p* = .550, *d* = 0.136; right: *t*(228) = 1.423, *p* = .156, *d* = 0.322). For younger participants, we also found a significant fixed effect of hemisphere on LC contrast (*p* < .001, *d* = 1.462), with left LC contrast being higher than right LC contrast, but no other fixed effects were significant. For older participants (Table 2B), we did not find a significant training condition x timepoint interaction (*p* = .713, *d* = 0.073), but we did observe a significant fixed effect of timepoint (*p* = .046, *d* = 0.400), with LC contrast being higher at the post-compared to the pre-training timepoint. As in the younger sample, we observed a significant effect of hemisphere on LC contrast in older participants (*p* < .001, *d* = 1.261), driven by higher contrast for the left relative to the right LC. Notably, in a model including data from both age groups, we did not observe significant timepoint x condition, age x timepoint x condition, or age x timepoint interactions on LC contrast (*p*’s > .05; Supplementary Results, Section 2).

**Figure 2.**
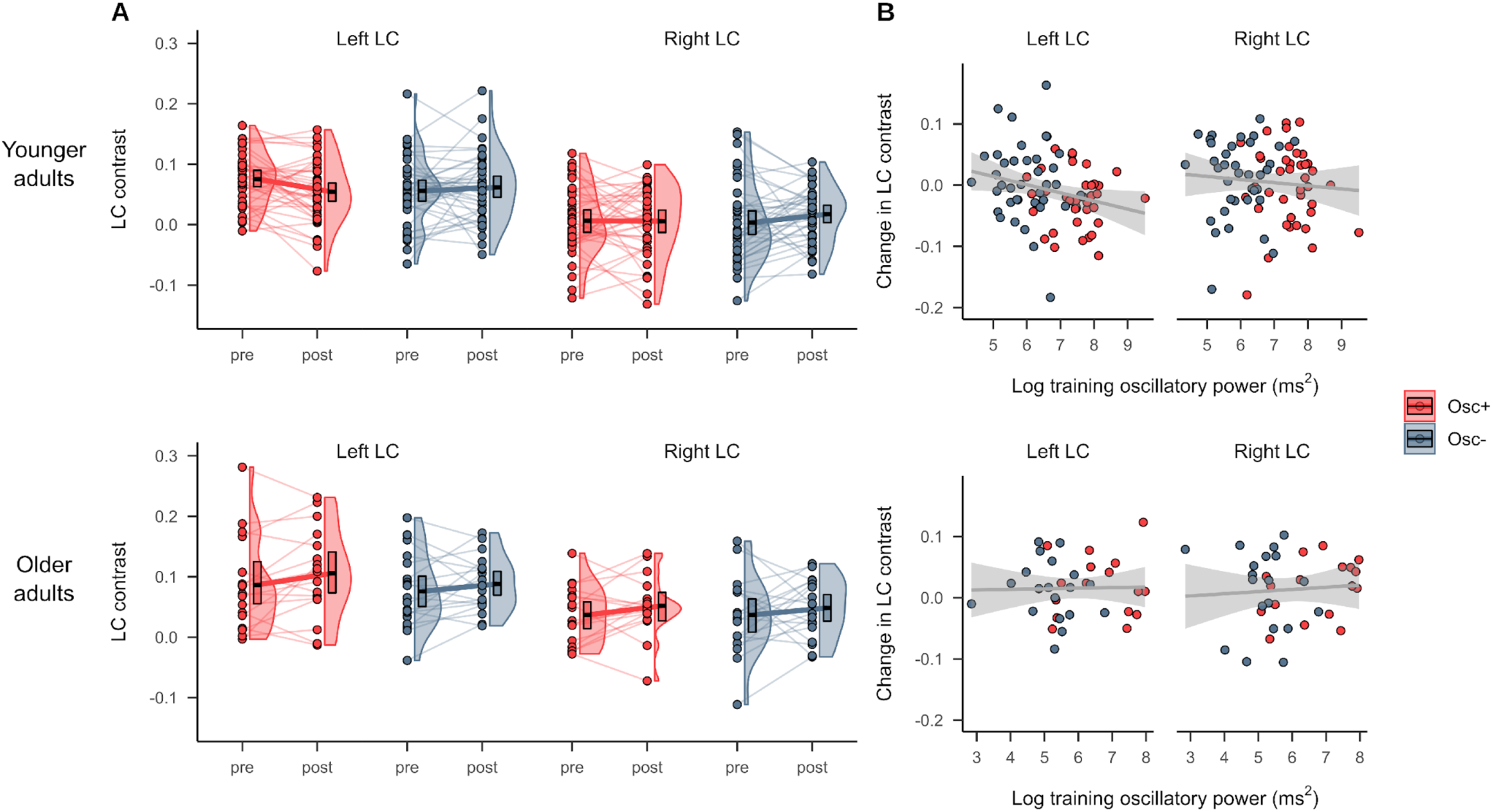
(A) LC MRI contrast at the pre- and post-training timepoints for the Osc+ and Osc-conditions, for younger (top) and older (bottom) participants. (B) Associations between pre- to post-training change in LC contrast and training oscillatory power, a measure of how much participants increased their heart rate oscillations across practice sessions, for younger (top) and older (bottom) participants. Linear regression lines with 95% confidence intervals are shown in gray.

**Table 2:**
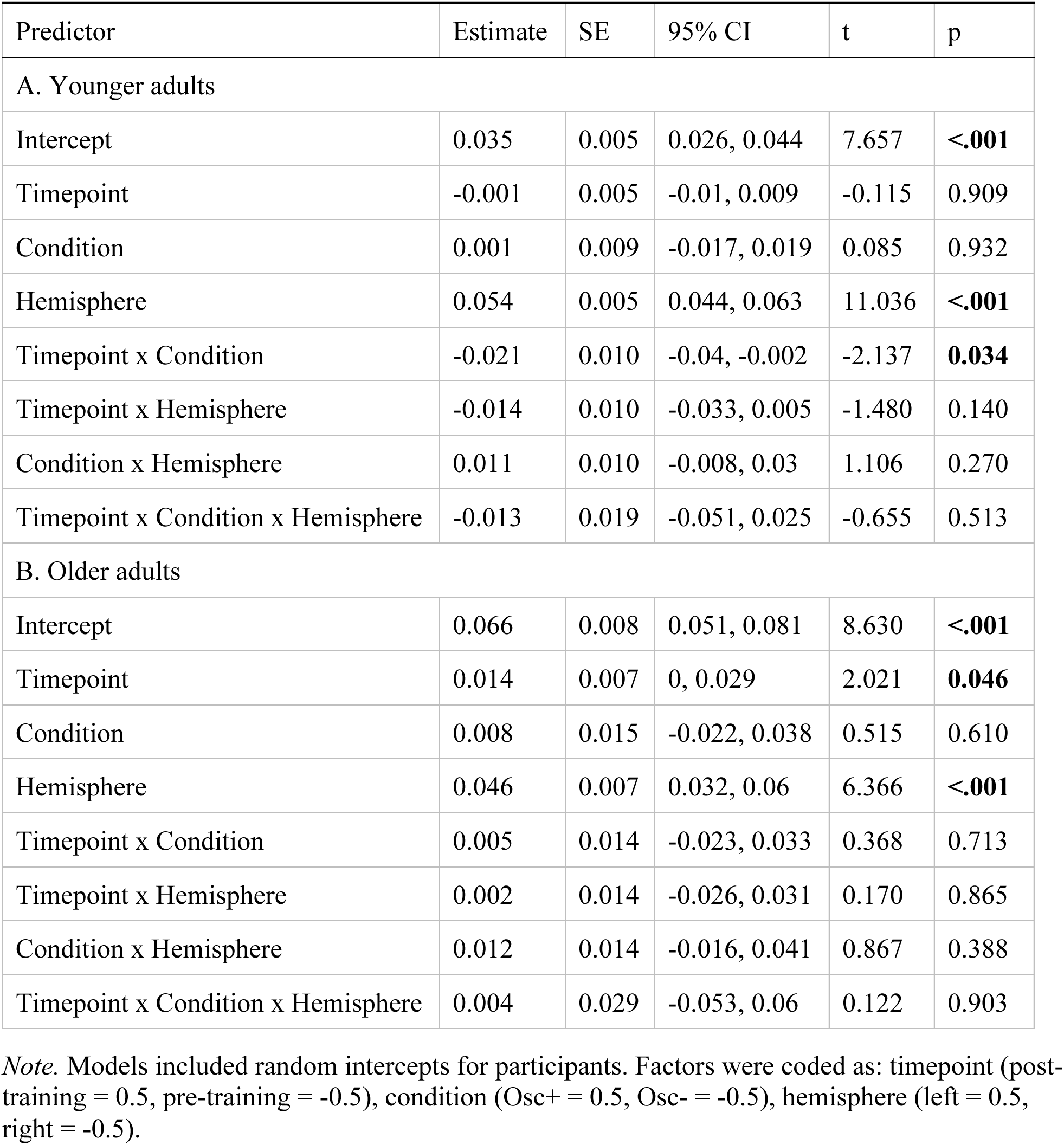
Results of linear mixed effects analysis testing the fixed effects of timepoint, training condition, and hemisphere on LC contrast, in younger adults (A) and older adults (B).

| Predictor | Estimate | SE | 95% CI | t | p |
| --- | --- | --- | --- | --- | --- |
| A. Younger adults |  |  |  |  |  |
| Intercept | 0.035 | 0.005 | 0.026, 0.044 | 7.657 | **<.001** |
| Timepoint | -0.001 | 0.005 | -0.01, 0.009 | -0.115 | 0.909 |
| Condition | 0.001 | 0.009 | -0.017, 0.019 | 0.085 | 0.932 |
| Hemisphere | 0.054 | 0.005 | 0.044, 0.063 | 11.036 | **<.001** |
| Timepoint x Condition | -0.021 | 0.010 | -0.04, -0.002 | -2.137 | **0.034** |
| Timepoint x Hemisphere | -0.014 | 0.010 | -0.033, 0.005 | -1.480 | 0.140 |
| Condition x Hemisphere | 0.011 | 0.010 | -0.008, 0.03 | 1.106 | 0.270 |
| Timepoint x Condition x Hemisphere | -0.013 | 0.019 | -0.051, 0.025 | -0.655 | 0.513 |
| B. Older adults |  |  |  |  |  |
| Intercept | 0.066 | 0.008 | 0.051, 0.081 | 8.630 | **<.001** |
| Timepoint | 0.014 | 0.007 | 0, 0.029 | 2.021 | **0.046** |
| Condition | 0.008 | 0.015 | -0.022, 0.038 | 0.515 | 0.610 |
| Hemisphere | 0.046 | 0.007 | 0.032, 0.06 | 6.366 | **<.001** |
| Timepoint x Condition | 0.005 | 0.014 | -0.023, 0.033 | 0.368 | 0.713 |
| Timepoint x Hemisphere | 0.002 | 0.014 | -0.026, 0.031 | 0.170 | 0.865 |
| Condition x Hemisphere | 0.012 | 0.014 | -0.016, 0.041 | 0.867 | 0.388 |
| Timepoint x Condition x Hemisphere | 0.004 | 0.029 | -0.053, 0.06 | 0.122 | 0.903 |

*Note.* Models included random intercepts for participants. Factors were coded as: timepoint (post-training = 0.5, pre-training = -0.5), condition (Osc+ = 0.5, Osc- = -0.5), hemisphere (left = 0.5, right = -0.5).

### 3.2 Training oscillatory power was associated with decreases in left LC contrast

Associations between training oscillatory power and change in LC contrast are depicted in Figure 2B. In younger participants, we found a significant negative correlation between training oscillatory power and change in left LC contrast (*r*(74) = -0.249, *95% CI* = -0.449 - -0.025, *p* = .030) but no significant correlation between training oscillatory power and change in right LC contrast (*r*(74) = -0.085, *95% CI* = -0.305 - 0.143, *p* = .463). In older participants, training power was not correlated with change in either left LC contrast (*r*(34) = 0.024, *95% CI* = -0.307 - 0.35, *p* = .889) or right LC contrast (*r*(34) = 0.076, *95% CI* = -0.259 - 0.395, *p* = .660). We note that the negative association between training power and left LC contrast change in younger adults did not emerge in a linear mixed effects analysis testing the fixed effects of training power, age group, hemisphere and their interactions on LC contrast; specifically, this analysis indicated no significant fixed effects of training power or interaction effects involving training power (*p*’s > .05; Supplementary Results, Section 3).

### 3.3 The association between training oscillatory power and change in left LC contrast was more negative in males

Pre- and post-training LC contrast for younger males and females is shown in Figure 3A. A linear mixed effects analysis testing fixed effects of timepoint, condition, hemisphere, sex and their interactions in younger adults indicated no significant timepoint x condition x sex or timepoint x condition x hemisphere x sex interactions on LC contrast (*p*’s > 0.05; Supplementary Results, Section 4). We note that this analysis indicated a significant fixed effect of sex on LC contrast (*p* = .007, *d* = 0.650), driven by greater LC contrast for females than males, as well as a significant timepoint x condition interaction on LC contrast (*p* = .032, *d* = -0.290), in line with what we observed above (Section 3.1). When we next added sex as a fixed effect to the model testing the effects of training oscillatory power and hemisphere on change in LC contrast (Table 3), we found a marginally significant interaction between training power and sex (*p* = .050, *d* = 0.470). This was driven by younger males having a more negative association between training power and change in LC contrast than younger females (Figure 3B). Notably, this analysis also indicated a significant fixed effect of sex (*p* = .048, *d* = -0.473), with females exhibiting greater decreases in LC contrast relative to males, and a significant fixed effect of training oscillatory power on change in LC contrast (*p* = .036, *d*= -0.505), after accounting for the effects of sex and hemisphere.

**Figure 3.**
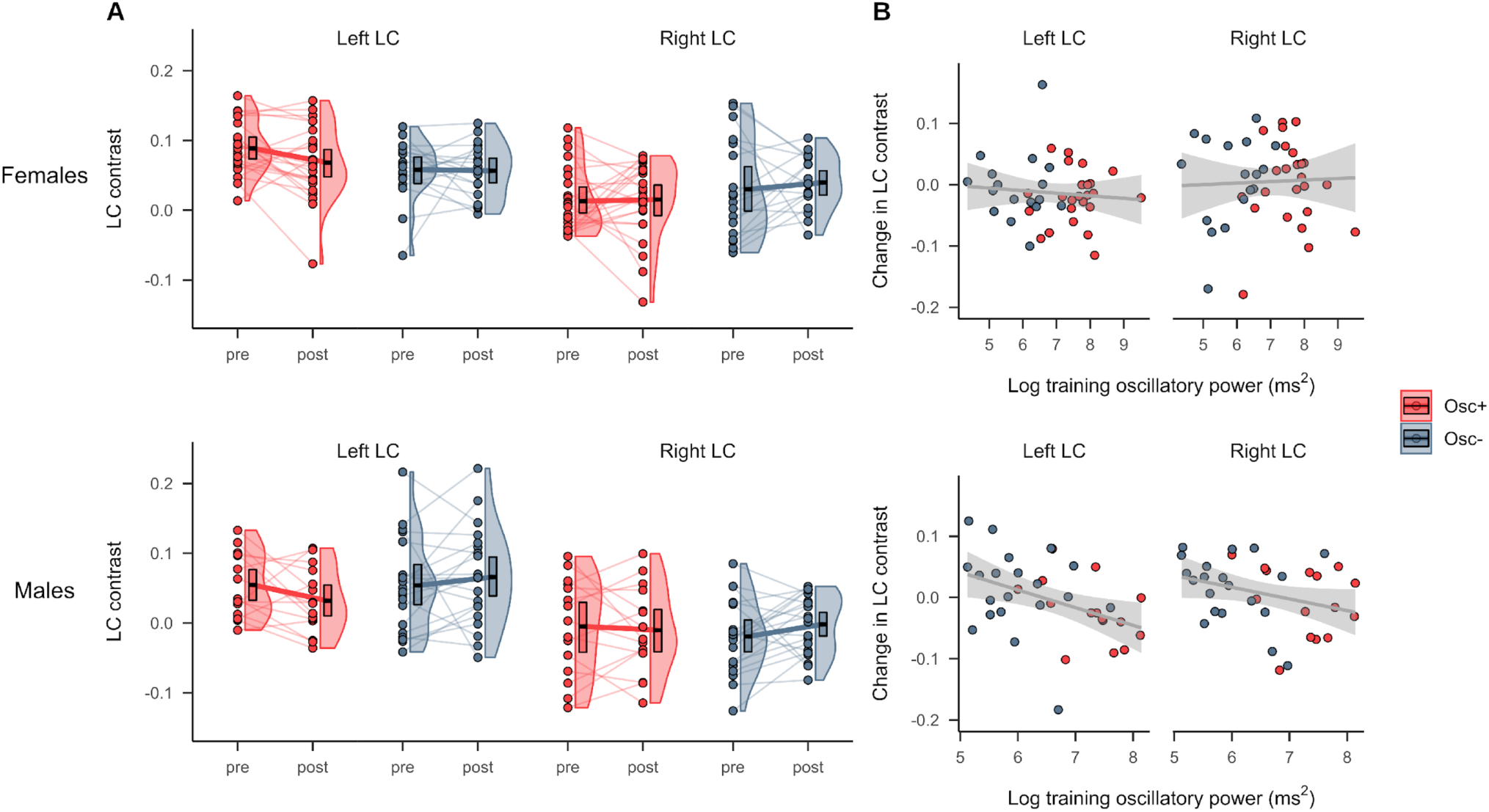
(A) LC MRI contrast at the pre- and post-training timepoints for younger participants in the Osc+ and Osc-conditions, stratified by sex (top = females, bottom = males). (B) Associations between change in LC contrast and training oscillatory power among younger participants, stratified by sex (top = females, bottom = males). Linear regression lines with 95% confidence intervals are shown in gray.

**Table 3:** Results of linear mixed effects analysis testing the fixed effects of training oscillatory power, hemisphere, and sex on change in LC MRI contrast.

| Predictor | Estimate | SE | 95% CI | t | p |
| --- | --- | --- | --- | --- | --- |
| Intercept | 0.080 | 0.039 | 0.004, 0.156 | 2.050 | **0.044** |
| Training power | -0.012 | 0.006 | -0.024, -0.001 | -2.142 | **0.036** |
| Hemisphere | 0.041 | 0.049 | -0.055, 0.137 | 0.838 | 0.405 |
| Sex | -0.157 | 0.078 | -0.309, -0.004 | -2.008 | **0.048** |
| Training power x Hemisphere | -0.008 | 0.007 | -0.022, 0.006 | -1.132 | 0.261 |
| Training power x Sex | 0.023 | 0.012 | 0, 0.046 | 1.993 | **0.050** |
| Hemisphere x Sex | -0.027 | 0.098 | -0.218, 0.165 | -0.273 | 0.785 |
| Training power x Hemisphere x Sex | 0.003 | 0.015 | -0.025, 0.032 | 0.215 | 0.830 |

*Note.* Models included random intercepts for participants. Factors were coded as: hemisphere (left = 0.5, right = -0.5), sex (female = 0.5, male = -0.5).

### 3.4 Decreases in left LC contrast were associated with decreases in CREB activity

Results of RNA sequencing in younger participants with available blood-based measures indicated a significant interaction between timepoint and condition on expression of genes regulated by the SNS-responsive CREB transcription factor (*bootstrap z* = -3.30, *p* = .001. Younger participants in the Osc-condition showed what appears to be a secular trend, with increased CREB activity from pre- to post-training (*z* = 2.70, *p* = .008), whereas participants in the Osc+ condition were buffered against that trend, showing no significant change over time (*z* = -0.45, *p* = .650).

We also found that greater change in LC contrast was associated with greater change in CREB activity (Figure 4), selectively for left LC contrast (*z* = 1.97, *p* = .049), with no significant effect for right LC contrast (*z* = 0.63, *p* = .530). In other words, participants with larger decreases in left LC contrast had larger decreases in CREB activity.

**Figure 4.**
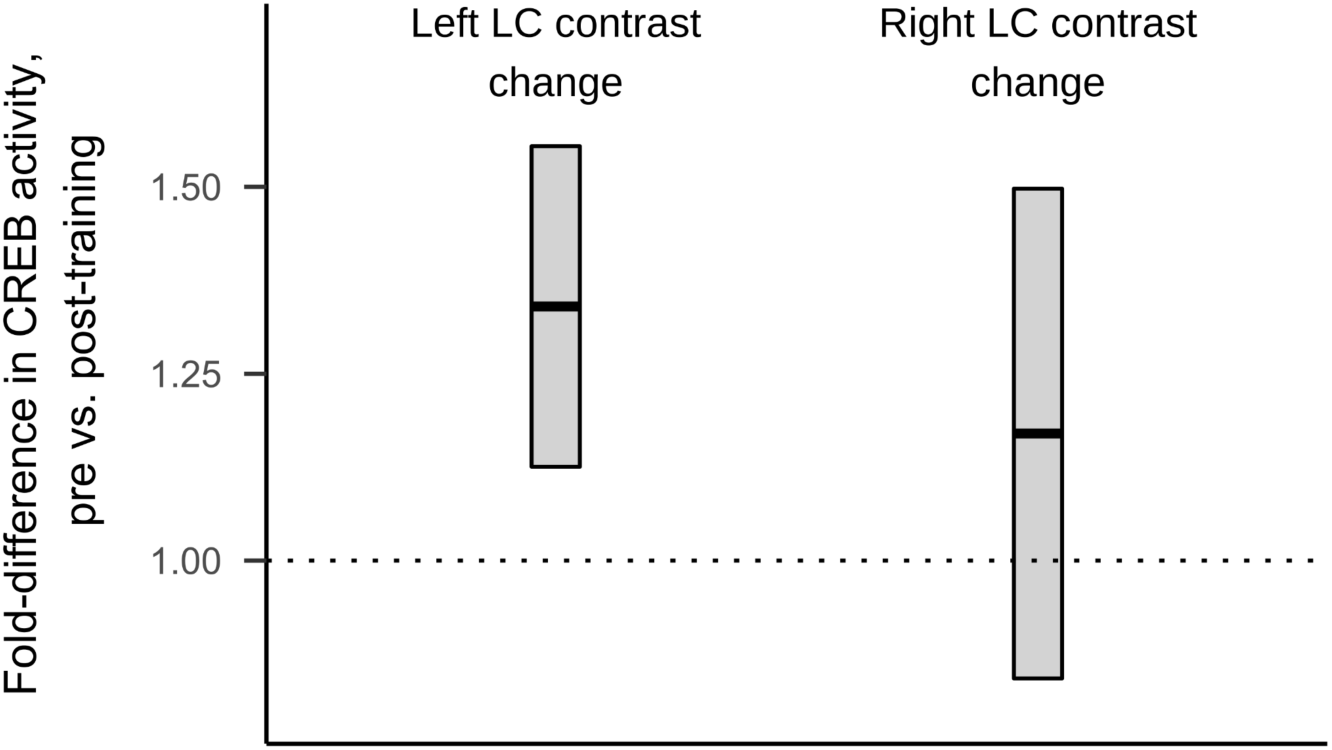
Fold-difference metrics reflecting pre- to post-training elevation in CREB activity in genes that showed >1.5 fold-differential expression per standard deviation of left and right LC contrast change. Crossbar central lines indicate the mean fold-differences, with a mean fold-difference of 1 corresponding to no pre- to post-training difference in CREB activity. Upper and lower bounds of crossbars extend reflect standard errors derived from bootstrap resampling of linear model residual vectors in underlying gene expression data. Figure reflects data from a subset of 39 younger participants for whom both blood-based measures and LC contrast values were available.

## 4 Discussion

In recent years, much has been learned about how LC MRI contrast, a proxy for LC structural integrity, relates to cognition across the lifespan (Betts et al., 2019; Elman et al., 2021; Liu et al., 2020). However, no published studies have examined factors that influence LC contrast across time. Here, we found that in younger adults, performing 5 weeks of HRV biofeedback training decreased LC contrast. This effect was larger for the left LC and scaled with the extent to which participants increased their heart rate oscillations during training. We also found that among younger participants with available blood-based measures, decreases in left LC contrast were coupled with decreases in activity of the CREB transcription factor that mediates SNS signaling through beta-adrenergic receptors (Cole et al., 2010; Mayr & Montminy, 2001). On the contrary, among older adults who completed biofeedback training, we did not observe training effects on LC contrast. Thus, for younger adults, using biofeedback to increase heart rate oscillations in daily training sessions affected LC MRI contrast.

Why might HRV biofeedback training have decreased LC contrast in younger adults? The beneficial effects of HRV biofeedback involving slow breathing are thought to occur through multiple mechanisms, including stimulation of the vagus nerve (Huang et al., 2018; Lehrer & Gevirtz, 2014). The vagus nerve is a major component of the parasympathetic nervous system and sends inputs to the LC via the medullary NTS (Badran et al., 2018; Fornai et al., 2011). The NTS is affected by respiration, with NTS cell firing suppressed during inhalation and facilitated during exhalation (Miyazaki et al., 1998). Thus, the balance of respiratory phases may affect LC activity. In support of this idea, exhalation-gated auricular vagal afferent nerve stimulation elicits greater responses in NTS and LC compared to inhalation-gated stimulation in humans (Garcia et al., 2017; Sclocco et al., 2019). One possibility is thus that the repeated practice of slow, paced breathing leads to more phasic and less tonic stimulation of the NTS and LC. In addition, a cluster of neurons in the medullary preBötzinger complex serves as a major breathing rhythm generator and provides excitatory input to the LC; when breathing is slow, the preBötzinger cluster provides less excitatory input to the LC, promoting lower tonic levels of arousal (Yackle et al., 2017). These slow-breathing effects on the NTS and preBötzinger neurons would have the net effect of shifting LC activity to a higher phasic and lower tonic level, which would manifest as lower LC MRI signal contrast and reduced sympathetic activity. The association we observed between decreases in LC contrast and decreases in activity of the CREB transcription factor are consistent with the notion of decreased LC contrast in younger adults reflecting decreased cumulative noradrenergic activity during the intervention time frame.

More broadly, our effects may be accounted for by an overall shift to parasympathetic dominance that occurs with the repeated practice of HRV biofeedback training. The LC receives projections from the medulla’s nucleus paragigantocellularis (Aston-Jones et al., 1986; Aston-Jones et al., 1991), which itself receives widespread autonomic inputs and has been implicated in the regulation and control of sympathetic activity and respiration (Van Bockstaele & Aston-Jones, 1995). Parasympathetic/sympathetic balance is then expected to directly impact the LC. As correlational evidence for this idea in humans, we previously found that HRV was negatively associated with LC MRI contrast in younger adults (Mather et al., 2017). In addition, LC efferent projections provide excitatory control over preganglionic sympathetic neurons and inhibitory control over the parasympathetic dorsal motor vagal nucleus and nucleus ambiguus (Samuels & Szabadi, 2008). Having relatively lower LC structural integrity would therefore give rise to less excitatory input to sympathetic centers and reduced inhibition of parasympathetic centers, as well as reduced excitatory input to the central nucleus of the amygdala by LC neurons, which also contribute to sympathetic activation (Wood et al., 2017).

We found that the effects of biofeedback were larger for the left than the right LC. Decreases in LC contrast for participants in the Osc+ condition were greater for the left than the right LC, and significant associations with training oscillatory power and CREB activity were observed for the left, but not the right, LC. Previous studies have reported higher MRI-assessed LC integrity in the left compared to the right LC (Betts et al., 2017; Dahl et al., 2019; Liu et al., 2019). Our findings are also in line with reports of more positive associations between LC contrast and cortical thickness for left relative to right LC (Bachman et al., 2021), as well as hemispheric differences in functional connectivity of the LC (Jacobs et al., 2018).

We also observed sex differences in how training oscillatory power related to change in LC contrast among younger participants, with males exhibiting a more negative association between training power and change in LC contrast than females. Relative to that of males, the female LC exhibits morphological and functional differences: LC neurons are more sensitive to corticotropin releasing factor (Bangasser et al., 2016) and exhibit greater dendritic density and branching (Bangasser et al., 2011; Ross & Van Bockstaele, 2020) in females. In line with previous reports of higher MRI-assessed measures of LC integrity in females than males (Bachman et al., 2021; Riphagen et al., 2020), we found that younger females had higher LC contrast than males across conditions, hemispheres and timepoints. Our findings of change being more coupled with training oscillatory power in younger males than females suggests that there are sex differences in the factors that shape LC contrast over time, which warrants further investigation.

Although we observed differential effects of the two HRV biofeedback training conditions on LC contrast among younger participants, this was not the case among older participants. Instead, among older participants, there was an overall increase in LC contrast from pre- to post-training. One possibility is that our study was not sufficiently powered to detect condition-specific effects in older adults; data collection for the older cohort was terminated early due to the COVID-19 pandemic. Another possibility is that among older adults, LC contrast reflects neurodegeneration more so than stress, whereas the opposite is true in younger adults. This means that an intervention affecting the autonomic nervous system would be more likely to change younger adults’ LC contrast levels than older adults’. Finally, the similar changes seen across the two conditions raise the possibility that older adults’ LC contrast levels were sensitive to an aspect of the intervention not explored here and present in both training conditions.

There are several other limitations to note. First, RNA sequencing analyses included only a subset of younger participants as we started collecting blood samples after some participants had completed the study. Second, participants in this study included mostly university students, limiting the external validity of results and potentially introducing a secular trend towards greater SNS activity as the 7-week study progressed; we aimed to avoid semester breaks in the study and therefore, across conditions, enrolled most younger participants at the beginning of semesters, when there are usually fewer exams and deadlines relative to later weeks in the semester. Third, our study only encompassed 5 weeks of HRV biofeedback training, but training over longer time periods may yield larger effects on LC contrast in both hemispheres and in younger and older participants. Finally, a limitation to the study is our limited understanding of the stability of the LC contrast measure over time. Our findings would be better contextualized by future studies which assess the stability of LC contrast in younger and older adults over time.

In this study, we assessed the effect of performing 5 weeks of heart rate variability biofeedback training on LC contrast, a measure that has been linked to cognition in older adults and arousal and negative affect in younger adults. We found that training decreased left LC contrast among younger participants and this effect scaled with the extent to which participants increased their heart rate oscillations during training. Furthermore, decreases in left LC contrast were related to decreases in CREB activity, a marker of sympathetic nervous system activity. These results provide novel evidence that among younger adults, LC contrast can be changed through the daily practice of increasing heart rate oscillations.

## Data Availability

Data supporting the findings of this study will be made publicly available at OpenNeuro.

## Acknowledgments

We are grateful to Sumedha Attanti and Juliana Lee who performed manual LC delineation and the following individuals who helped collect data for the project: Collin Amano, Kathryn Cassutt, Christine Cho, Akanksha Jain, Katherine Jeung, Sophia Ling, Althea Wolfe, Michelle Wong, Yong Zhang, and Cyndy Zhuang. We also thank Martin Dahl for advice on LC delineation.

This work was supported by National Science Foundation grant number DGE-1842487, and by the National Institute on Aging of the National Institutes of Health under award numbers R01AG057184, P30AG017265, and T32AG000037. The content is solely the responsibility of the authors and does not necessarily represent the official views of the National Institutes of Health.

