## Supplementary Material for "Daily heart rate variability biofeedback training decreases locus coeruleus MRI contrast in younger adults"

### Supplementary Methods

#### Section 1. Details of included and excluded TSE scans

A total of 115 younger (60 Osc+, 55 Osc-) and 60 older (31 Osc+, 29 Osc-) participants pre- and/or post-training MRI assessments including a TSE scan. Of these, 15 younger and 9 older participants did not finish the study and thus did not complete a post-training MRI assessment. The proportion of participants who did not complete the study was higher for older than for younger adults because we terminated the study early due to the COVID-19 pandemic; at that point, 6 older participants were unable to complete the study. There was 1 older adult participant whose pre-training MRI assessment did not include a TSE scan. A breakdown of MRI assessments that included MPRAGE and TSE scans, according to age group, training condition and timepoint, is provided in Table S1.

Table S1:

*Description of MRI assessments that included MPRAGE and TSE scans.*

| Age group | Training condition | Timepoint | Number of assessments |
|---|---|---|---|
| Younger | Osc+ | Pre-training | 60 |
| Younger | Osc+ | Post-training | 52 |
| Younger | Osc- | Pre-training | 55 |
| Younger | Osc- | Post-training | 48 |
| Older | Osc+ | Pre-training | 31 |
| Older | Osc+ | Post-training | 26 |
| Older | Osc- | Pre-training | 28 |
| Older | Osc- | Post-training | 25 |

*Note.* Osc+ = increase-oscillations condition; Osc- = decrease-oscillations condition.

Available TSE scans were visually inspected by one trained researcher for quality and artifact. Scans with incorrect positioning (n = 3), different resolution (n = 1), or susceptibility artifact overlapping the LC or central pons (n = 5) were excluded from LC delineation. Of remaining scans, 29 were excluded due to excessive motion. As a validation step, we compared whether scans were included or excluded with qualitative information provided by two raters during manual LC delineation (Section 3); specifically, in some cases, one or both raters reported not being able to manually identify or delineate the LC due to artifact. Scans that could not be rated by either rater were considered non-rateable, whereas scans rated by at least one rater were considered rateable. We then found 93.7% agreement between whether scans had been flagged as included or excluded based on visual inspection, and their rateability. Because raters were told to prioritize delineating the LC even if the surrounding image contained excessive motion or artifact, inclusions and exclusions from visual inspection were taken as final decisions.

**Section 2. LC delineation parameters and validation**

We first upsampled available MPRAGE and FSE scans to twice their native resolution. For this step, the ResampleImage ANTs routine was used, using linear interpolation and pixel type set to 'float.'

We then generated an initial, whole-brain template using a subset of 134 upsampled MPRAGE scans whose fields of view were spatially well-aligned, specifically with `qoffset_x`, `qoffset_y`, and `qoffset_z` values falling within 1 standard deviation of the mean across scans. The subset included scans from 88 younger and 46 older participants. These scans were used as inputs to a run of the antsMultivariateTemplateConstruction.sh routine with the following parameters: gradient step size = 0.25, iteration limit = 6, max iterations = 1x0x0, modality weights used in similarity metric = 1, number of modalities = 1, N4 bias field correction on, rigid body registration of inputs on, registration similarity metric = cross-correlation, transformation model type = greedy-SyN, update template with full affine transform on, no initial template. The resulting template was used as the initial template for a full template-building run of the antsMultivariateTemplateConstruction.sh routine, using all 287 (191 younger, 96 older) MPRAGE scans as inputs. This template-building run had the same parameters as the initial template-building run, except for the following: max iterations = 30x90x20, rigid body registration of inputs off. The result was a full, whole-brain (MPRAGE) template, as well as each inputted MPRAGE scan coregistered to whole-brain template space.

Coregistration of upsampled TSE scans to corresponding whole-brain template-coregistered MPRAGE scans was performed using antsRegistrationSyNQuick.sh, with the following parameters: transform type = rigid, affine and deformable SyN ('s'), histogram bins = 32, spline distance = 26, precision type = double, transform type = SyN, histogram matching on. Coregistered TSE scans were then used to create a TSE template. Parameters for this template-building procedure were the same as that for the whole-brain template building procedure, except that all coregistered TSE scans were used for both the initial and full template-building runs.

We then coregistered the full TSE template with the whole-brain template, and we coregistered the whole-brain template with the MNI152 0.5mm (linear) brain. For both coregistration steps, the antsRegistrationSyN.sh routine was used with the following parameters: transform type = rigid, affine and deformable SyN ('s'), radius = 4, spline_distance = 26, precision type = double, histogram matching on, collapse output transforms on. For coregistration of the TSE template to MPRAGE template space, a binarized lesion mask was used which was constructed by thresholding the TSE template at intensity 1. Finally, using transforms from all steps above, we warped upsampled TSE scans to MNI152 0.5mm (linear) space, using the antsApplyTransforms.sh routine, with interpolation type set to linear. The antsApplyTransforms.sh, with the same parameters, was used to apply relevant transforms to warp the TSE template to MNI152 space.

As a validation step, hyperintensities on the TSE template that had been warped to MNI152 space were compared with the locations of a publicly available LC meta-map (Dahl et al., 2021). These hyperintensities are displayed alongside the meta-map coordinates in Figure S1.

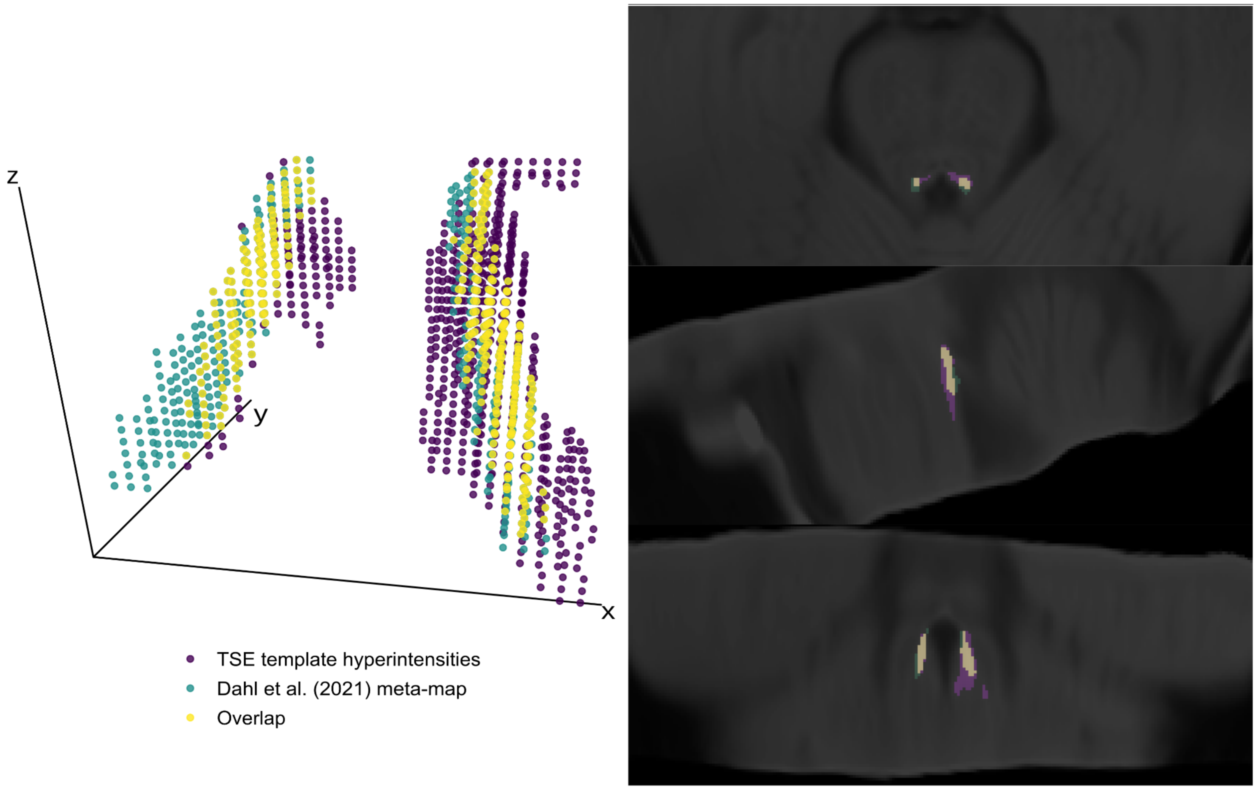


*Figure S1.* Comparison of signal hyperintensity locations on the TSE template, LC meta-map from Dahl et al. (2021), and their overlap. The TSE template was warped to MNI152 0.5mm (linear) standard space. Locations of hyperintensity were all voxels in the dorsal pons that survived thresholding based on intensities within the central pontine reference region. Specifically, the Dahl et al. (2021) reference map was applied as a mask to the TSE template. For each slice in the *z*-direction ($z$= 85-112), an intensity threshold was computed as the mean reference intensity in that slice plus 3.5 times the standard deviation of the reference intensity in that slice. Then, voxels within the same slice of the dorsal pons ($x$ = 164-196, $y$ = 174-182) where intensities were greater than the threshold value were classified as hyperintensities. Left panel shows graphical comparison of meta-mask, template hyperintensities, and their overlap. Right panel shows the same comparison overlaid on the TSE template in MNI space (x = 187, y = 179, z = 98).

**Section 3. Manual LC delineation**

As a validation step, peak LC locations were also manually delineated on native-resolution TSE scans by two trained raters. Raters were blind to training condition, age and timepoint. Manual LC delineation was performed using ImageJ (Version 1.5.2, Schneider et al., 2012, https://imagej.nih.gov). according to the protocol described by Dahl et al. (2019). In summary, raters identified 2x2 voxel regions of interest in the left and right hemispheres which exhibited peak intensity and overlapped the expected location of the LC. This procedure was performed by each rater for each *z*-slice in which the left and/or right LC was visible.

For each rater, the voxel with peak LC intensity was then selected across *z*-slices, for each hemisphere separately. We then used two-way mixed-effects intra-class correlation analyses to assess correspondence between peak left and right LC intensity values across raters. These analyses indicated high correspondence between raters for the left LC (*ICC*(A, 1) = 0.966, *95%*

*CI* = 0.951 - 0.975, $p < .001$) and right LC (*ICC*(A, 1) = 0.968, *95% CI* =0.958 - 0.976, $p < .001$). Therefore, we averaged left and right LC intensity values across raters. The resulting averaged values were used to assess correspondence between intensity values determined manually and using the semi-automated method (main text, Section 2.3.1).

**Section 4. Training pulse data collection and calculation of training oscillatory power**

During each biofeedback training session, pulse was measured using HeartMath emWave Pro software with an infrared pulse plethysmograph ear sensor with a sampling rate of 370 Hz. Interbeat interval data for each session were extracted from pulse data after the elimination of ectopic beats or other sources of artifact through a built-in process in emWave Pro.

As a measure of how much each participant increased their heart rate oscillations on average during biofeedback training, we computed a measure of training oscillatory power as follows. Kubios HRV Premium (Version 3.1, Tarvainen et al., 2014, https://www.kubios.com/hrv-premium/) was used to calculate autoregressive spectral power from the interbeat interval data from each biofeedback training session. The following parameters were applied during analysis with Kubios: automatic artifact correction, 500-lambda smoothing priors detrending method, autoregressive order = 16, no autoregressive factorization, and all other parameters set to default values. For each training data segment, we summed power values within the frequency range from 0.063 - 0.0125 Hz, which corresponded to the range of participants' potential breathing paces (8-16s per breath). A single value of training power for each participant was computed by averaging across power values from all training sessions. Power values were log transformed prior to statistical analysis.

**Section 5. Testing for sex differences in effects on LC contrast**

To test for sex differences in how HRV biofeedback training affected LC contrast in younger adults, we used a linear mixed effects analysis, specifying fixed effects of training condition, timepoint, hemisphere, sex and their interactions and LC contrast as the dependent variable. To test for sex differences in the association between LC contrast change and training oscillatory power, we performed another analysis, specifying fixed effects of training power, hemisphere, sex, and their interactions and change in LC contrast as the dependent variable. For these analyses, sex was coded as female = 0.5, male = -0.5.

### Supplementary Results

#### Section 1. Analysis of distance between pre- and post-training peak LC locations

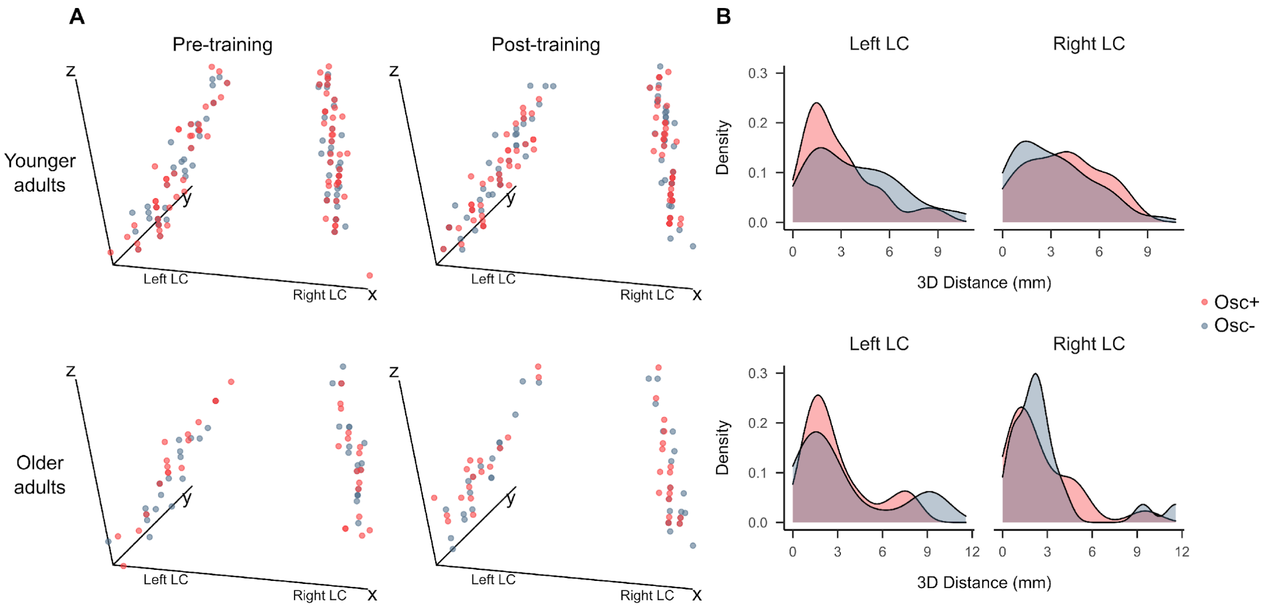


*Figure S2.* (A) Coordinates of peak LC signal intensity at the pre- and post-training timepoints in MNI152 0.5mm linear standard space. Coordinates range from x = 192 (left) to 168 (right), y = 172 (front) to 182 (back), z = 85 (bottom) to 112 (top). (B) Density plots depicting 3-dimensional distance in millimeters between each participant's peak LC intensity coordinates at the pre- and post-training timepoints. Osc+ = increase-oscillations condition; Osc- = decrease-oscillations condition.

Table S2:

*Results of a linear mixed effects analysis testing the fixed effects of training condition, age group and hemisphere on 3-dimensional distance between pre- and post-training peak LC intensity locations.*

| Predictor | Estimate | SE | 95% CI | t | p |
|---|---|---|---|---|---|
| Intercept | 1.648 | 0.052 | 1.545, 1.75 | 31.513 | **<.001** |
| Condition | -0.050 | 0.105 | -0.255, 0.155 | -0.476 | 0.635 |
| Hemisphere | 0.048 | 0.095 | -0.139, 0.235 | 0.507 | 0.613 |
| Age group | -0.144 | 0.105 | -0.349, 0.061 | -1.380 | 0.170 |
| Condition x Hemisphere | -0.186 | 0.191 | -0.559, 0.188 | -0.975 | 0.332 |
| Condition x Age group | 0.001 | 0.209 | -0.409, 0.411 | 0.006 | 0.995 |
| Hemisphere x Age group | 0.133 | 0.191 | -0.241, 0.506 | 0.697 | 0.488 |
| Condition x Hemisphere x Age group | 0.500 | 0.381 | -0.247, 1.247 | 1.311 | 0.193 |

*Note.* Model included random intercepts for participants. Factors were coded as: condition (Osc+ = 0.5, Osc- = -0.5), age group (older = 0.5, younger = -0.5), hemisphere (left = 0.5, right = -0.5). Distance values were square root transformed prior to analysis as a correction for non-normality, which was indicated by a significant Shapiro-Wilk test ($W = 0.904$, $p < .001$). The significant intercept term indicates that distances differed from 0 across conditions, age groups and hemispheres.

**Section 2. Analysis of training effects on LC contrast**

Table S3:

*Results of a linear mixed effects analysis testing the fixed effects of timepoint, training condition, age group and hemisphere on LC contrast.*

| Predictor | Estimate | SE | 95% CI | t | p |
|---|---|---|---|---|---|
| Intercept | 0.051 | 0.004 | 0.042, 0.059 | 11.903 | **<.001** |
| Timepoint | 0.007 | 0.004 | -0.002, 0.015 | 1.607 | 0.109 |
| Condition | 0.004 | 0.009 | -0.012, 0.021 | 0.511 | 0.611 |
| Hemisphere | 0.050 | 0.004 | 0.041, 0.058 | 11.463 | **<.001** |
| Age group | 0.031 | 0.009 | 0.015, 0.048 | 3.679 | **<.001** |
| Timepoint x Condition | -0.008 | 0.009 | -0.025, 0.009 | -0.895 | 0.371 |
| Timepoint x Hemisphere | -0.006 | 0.009 | -0.023, 0.011 | -0.690 | 0.491 |
| Condition x Hemisphere | 0.012 | 0.009 | -0.005, 0.029 | 1.338 | 0.182 |
| Timepoint x Age group | 0.015 | 0.009 | -0.002, 0.032 | 1.736 | 0.084 |
| Condition x Age group | 0.007 | 0.017 | -0.026, 0.04 | 0.419 | 0.676 |
| Hemisphere x Age group | -0.008 | 0.009 | -0.025, 0.009 | -0.931 | 0.353 |
| Timepoint x Condition x Hemisphere | -0.005 | 0.017 | -0.039, 0.029 | -0.267 | 0.790 |
| Timepoint x Condition x Age group | 0.026 | 0.017 | -0.008, 0.06 | 1.505 | 0.133 |
| Timepoint x Hemisphere x Age group | 0.017 | 0.017 | -0.017, 0.051 | 0.971 | 0.332 |
| Condition x Hemisphere x Age group | 0.002 | 0.017 | -0.032, 0.036 | 0.096 | 0.924 |
| Timepoint x Condition x Hemisphere x Age group | 0.016 | 0.035 | -0.052, 0.084 | 0.469 | 0.639 |

*Note.* Model included random intercepts for participants. Factors were coded as: timepoint (pre-training = 0.5, post-training = -0.5), condition (Osc+ = 0.5, Osc- = -0.5), age group (older = 0.5, younger = -0.5), hemisphere (left = 0.5, right = -0.5).

**Section 3. Analysis of associations between training power and change in LC contrast**

Table S4:

*Results of a linear mixed effects analysis testing fixed effects of training oscillatory power, age group and hemisphere on change in LC contrast.*

| Predictor | Estimate | SE | 95% CI | t | p |
|---|---|---|---|---|---|
| Intercept | 0.031 | 0.027 | -0.022, 0.083 | 1.147 | 0.254 |
| Training power | -0.003 | 0.004 | -0.012, 0.005 | -0.823 | 0.412 |
| Hemisphere | 0.029 | 0.035 | -0.04, 0.098 | 0.830 | 0.408 |
| Age group | -0.059 | 0.054 | -0.164, 0.046 | -1.104 | 0.272 |
| Training power x Hemisphere | -0.005 | 0.006 | -0.016, 0.006 | -0.970 | 0.334 |
| Training power x Age group | 0.011 | 0.008 | -0.005, 0.028 | 1.351 | 0.179 |
| Hemisphere x Age group | -0.023 | 0.071 | -0.162, 0.115 | -0.332 | 0.741 |
| Training power x Hemisphere x Age group | 0.006 | 0.011 | -0.016, 0.027 | 0.505 | 0.614 |

*Note.* Model included random intercepts for participants. Factors were coded as: age group (older = 0.5, younger = -0.5), hemisphere (left = 0.5, right = -0.5).

**Section 4. Analysis of sex differences in LC contrast change**

Table S5:

*Results of a linear mixed effects model testing the fixed effects of timepoint, training condition, hemisphere, and sex on LC contrast.*

| Predictor | Estimate | SE | 95% CI | t | p |
|---|---|---|---|---|---|
| Intercept | 0.034 | 0.004 | 0.025, 0.043 | 7.554 | **<.001** |
| Timepoint | -0.001 | 0.005 | -0.011, 0.008 | -0.218 | 0.828 |
| Condition | -0.003 | 0.009 | -0.021, 0.014 | -0.373 | 0.710 |
| Hemisphere | 0.052 | 0.005 | 0.042, 0.061 | 10.714 | **<.001** |
| Sex | 0.025 | 0.009 | 0.007, 0.042 | 2.794 | **0.007** |
| Timepoint x Condition | -0.021 | 0.010 | -0.04, -0.002 | -2.159 | **0.032** |
| Timepoint x Hemisphere | -0.014 | 0.010 | -0.033, 0.005 | -1.463 | 0.145 |
| Condition x Hemisphere | 0.011 | 0.010 | -0.008, 0.03 | 1.138 | 0.256 |
| Timepoint x Sex | -0.003 | 0.010 | -0.022, 0.016 | -0.306 | 0.760 |
| Condition x Sex | 0.007 | 0.018 | -0.028, 0.042 | 0.391 | 0.697 |
| Hemisphere x Sex | -0.017 | 0.010 | -0.036, 0.002 | -1.771 | 0.078 |
| Timepoint x Condition x Hemisphere | -0.012 | 0.019 | -0.05, 0.026 | -0.613 | 0.541 |
| Timepoint x Condition x Sex | 0.016 | 0.019 | -0.022, 0.054 | 0.821 | 0.412 |
| Timepoint x Hemisphere x Sex | -0.006 | 0.019 | -0.044, 0.032 | -0.290 | 0.772 |
| Condition x Hemisphere x Sex | 0.061 | 0.019 | 0.023, 0.099 | 3.165 | **0.002** |
| Timepoint x Condition x Hemisphere x Sex | 0.000 | 0.039 | -0.076, 0.076 | 0.010 | 0.992 |

*Note.* Model included random intercepts for participants. Factors were coded as: timepoint (pre-training = 0.5, post-training = -0.5), condition (Osc+ = 0.5, Osc- = -0.5), age group (older = 0.5, younger = -0.5), hemisphere (left = 0.5, right = -0.5), sex (female = 0.5, male = -0.5). Only data from younger adults was used for this analysis.
